# The Oncogenic Role of Human Microcephalin Gene Revealed by Pan-Cancer Analysis

**DOI:** 10.1101/2022.11.18.22282275

**Authors:** Zerui Wang, Mian Li, Zhen Liu, Weiming Kang, Yazhou Cui, Jinxiang Han, Wenbin Du

## Abstract

The human gene microcephalin (*MCPH1*) plays a key role in DNA damage-induced cellular responses and chromosome condensation. Recent clinical studies proposed *MCPH1* as a tumor suppressor gene in lung cancer, pancreatic cancer, and breast cancer, yet its roles remain poorly understood in other types of tumors. Pan-cancer analyses of *MCPH1* are urgently required to help us understand the potential molecular functions of *MCPH1* in other types of tumors. Here, we used several bioinformatic database and tools, including TCGA, GEO, ONCOMINE, and Human Protein Atlas to investigate the role of *MCPH1* in 33 tumor types. We found that the expression of *MCPH1* in tumor cases and normal cases were significantly different, and the higher expression of *MCPH1* generally predicted poor overall survival for tumor patients, such as acute myeloid leukemia, liver hepatocellular carcinoma, and pancreatic adenocarcinoma. Meanwhile, lower expression of the *MCPH1* gene was related to poor OS prognosis for KIRC and gastric cancer. Moreover, the expression level of *MCPH1* was highly associated with the immune microenvironment. Our result provides some fresh light into the oncogenic roles of *MCPH1* in various human cancers and revealed that *MCPH1* may be a potential diagnostic and prognostic marker in LAML, PAAD, and gastric cancer.

## Introduction

Cancer is the leading cause of death and a major burden of global public healthcare system. With the increasing cost of drug development and treatment for cancer, it has become increasingly important to predict potential marker genes and perform large-scale genomic analysis. Some of the published results of pan-cancer analysis have demonstrated significant contributions to clinical diagnosis and early treatment^1-4^. However, there are still many potential marker genes needed to be explored, and therefore comparative analyses of genes associated with cancer are urgently needed. What caught our attention was that microcephalin (*MCPH1*), a causative gene of primary autosomal recessive microcephaly 1, is low expressed in several malignancies^5-7^, yet there is no available research for *MCPH1* pan-cancer analysis.

Recently, *MCPH1* is believed to have played an important role in the origin and progression of several cancers. *MCPH1*, located on human chromosome 8p23.1, encodes for an 835 amino acid protein, containing several functional domains, such as regions that bind to condensin II, TopBP1, Chk1 and βTrCP2 and three BRCA1 C-terminus (BRCT) domains, and above-mentioned functional domains are generally found in proteins involved in DNA damage response and cell cycle regulation^8^. Evidence has been accrued that *MCPH1* is related to different malignancies. For example, the reduction of *MCPH1* was reported to be associated with BRCA1 negative breast cancer^9^, and MCPH1 protein expression and polymorphism were confirmed to be associated with breast cancer risk^10^. Liang et al. suggested that *MCPH1* deficiency promotes genomic instability and tumor formation in a mouse model^11^. In 2018, Wu et al. demonstrated that overexpression of *MCPH1* inhibits the migration and invasion of lung cancer cells^12^. These studies all highlight the potential of *MCPH1* as a cancer suppressor gene. However, there are few data-driven research related to effect of *MCPH1* in tumorigenesis, and it remains to be explored whether *MCPH1* is widely involved in the process of various tumor developments.

In this study, we performed a pan-cancer analysis of *MCPH1* across various cancers using the Cancer Genome Atlas (TCGA) and the Gene Expression Omnibus (GEO) datasets^13-15^. Out of these 33 cancers, 16 cancers had a significantly either higher or lower expression of *MCPH1* between normal and tumor tissues. Subsequently, we investigated the association between abnormal *MCPH1* expression and cancer prognosis. We next explored the relationship between the expression of *MCPH1* and immune microenvironment. Furthermore, we also investigated correlations between the expression of *MCPH1* and microsatellite instability (MSI) or tumor mutational burden (TMB). Our findings revealed a significant correlation between *MCPH1* and patient survival in gastric cancer, acute myeloid leukemia (LAML), and pancreatic adenocarcinoma (PAAD). These results suggest the potential value of MCPH1 as a biomarker of different types of cancer, which warrants further investigation.

## Methods

### Gene expression analysis

The ONCOMINE database (https://www.oncomine.org/)^16^, was used to analyze the mRNA expression of *MCPH1* in different cancer types. Next, we analyzed the expression profiling of *MCPH1* between tumors and adjacent normal tissues by the online tool TIMER2 (Tumor IMmune Estimation Resource, version 2, http://timer.cistrome.org/). For those tumors without or with highly limited normal tissues in the TCGA database, the GEPIA2 tool (http://gepia2.cancer-pku.cn/#analysis)^17^was used to analyze the box plots of expression differences between tumors and corresponding normal tissues of the Genotype-Tissue Expression (GTEx) database, under the settings of *P*-value cutoff = 0.01, log_2_FC (Fold change) cutoff = 1, and “Match TCGA normal and GTEx data”. GEPIA2 tool was used to analyze the *MCPH1* expression in different pathological stages (stage I, stage II, stage III, and stage IV) of all TCGA cancers. The log2 [TPM (Transcripts per million) + 1] transformed expression data were applied for the box or violin plots.

### Survival prognosis analysis

We used GEPIA2 to acquire the OS (Overall survival) and DFS (Disease-free survival) significance map data of *MCPH1* across all TCGA tumors. Cutoff-high (50%) and cutoff-low (50%) values were used as the expression thresholds for splitting the high-expression and low-expression cohorts. The survival plots were also obtained through the “Survival Analysis” module of GEPIA2^17^.

### Genetic alteration analysis

We used cBioPortal tool (https://www.cbioportal.org/)^18,19^ to obtain the data of mutation type, alteration frequency, mutated site information and Copy number alterations (CNAs) across all TCGA tumors. Additionally, survival data, including OS and DFS were compared for the TCGA cancer cases, with or without *MCPH1* genetic alteration.

### Immune microenvironment analysis

We used SangerBox tools, a free online platform for data analysis (http://vip.sangerbox.com/), to analyze the relationship between the expression of *MCPH1* and immune checkpoint genes, tumor purity and immune neoantigens. Pearson analysis was performed to assess the correlations between *MCPH1* and immune checkpoints, the results were displayed as heatmaps^20^. Immune and stromal score represented the abundance of immune and stromal components, respectively. ESTIMATE score was the sum of previous scores, representing tumor purity indirectly^20^. The correlations of *MCPH1* expression with these scores in different cancers were depicted as scatter plots.

Furthermore, we used TIMER2 tool^21,22^ to analyze the relationship between *MCPH1* expression and immune infiltrates across all TCGA tumors. The TIMER, TIDE, CIBERSORT, CIBERSORT-ABS, QUANTISEQ, XCELL, MCPCOUNTER and EPIC algorithms were applied for estimations.

### *MCPH1*-Related gene enrichment analysis

We used STRING (http://string-db.org/)^23^ to analyze the protein-protein interaction network. The main parameters were as follows: minimum required interaction score [“Low confidence (0.150)”], meaning of network edges (“evidence”), max number of interactors to show (“no more than 50 interactors” in 1st shell) and active interaction sources (“experiments”).

Based on the datasets of all tumors and normal tissues of TCGA, we used the GEPIA2 to get the top 100 *MCPH1*-correlated genes. Next, we performed a pairwise gene-gene Pearson correlation analysis of *MCPH1* and selected genes. Additionally, TIMER2 was used to calculate the heatmap data of selected genes, including the partial correlation (cor) and *p*-value in the purity-adjusted Spearman’s rank correlation test.

We further performed KEGG (Kyoto encyclopedia of genes and genomes) pathway analysis and GO (Gene ontology) enrichment analysis and visualized the results by the R language software [R-3.6.3, 64-bit] (http://www.r-project,org/).

### Ethical approval

All experimental protocols were conducted in accordance with relevant guidelines and regulations.

## Results

### Gene expression analysis

This study aims to explore the roles of *MCPH1* (NM_024596.5 for mRNA or NP_078872.3 for protein, **Fig. S1A**) in carcinogenesis in various human tumors. The conserved structural domains of *MCPH1* are Microcephalin (pfam 12258) and BRCT (cl00038). As can be seen in **Fig. S1B**, MCPH1 protein is relatively conserved in different species. The results of the developmental tree data (**Fig. S2**) also provide corresponding data to support the conserved nature of this protein.

Integrating the HPA (human protein atlas), GTEx, and FANTOM5 (functional annotation of the mammalian genome) databases, we analyzed the gene expression pattern of *MCPH1* in different cellular and non-tumor tissues, and the results were shown in **Fig. S3A**. *MCPH1* expression was highest in Bone marrow, followed by skeletal muscle and Thymus. However, *MCPH1* was detected in all tissues examined, showing a low RNA tissue specificity. We next explored the *MCPH1* expression in different blood cells, which revealed a low RNA blood cell specificity in **Fig. S3B**.

ONCOMINE database was used to explore the mRNA expression levels of *MCPH1* over a cancer-wide range. Lower *MCPH1* expression was found in breast, colorectal, lymphoma, and pancreatic cancer (**Fig. 1A**). However, our analysis showed higher *MCPH1* expression in other cancer groups, including breast, colorectal, and leukemia compared with normal groups. As shown in **Fig. 1B**, the expression of *MCPH1* was decreased in BLCA, BRCA, KICH, KIRC, KIRP, PRAD, READ, THCA, UCEC compared to normal tissues, and increased in some cancers such as CHOL, ESCA, STAD.

**Fig. 1.**
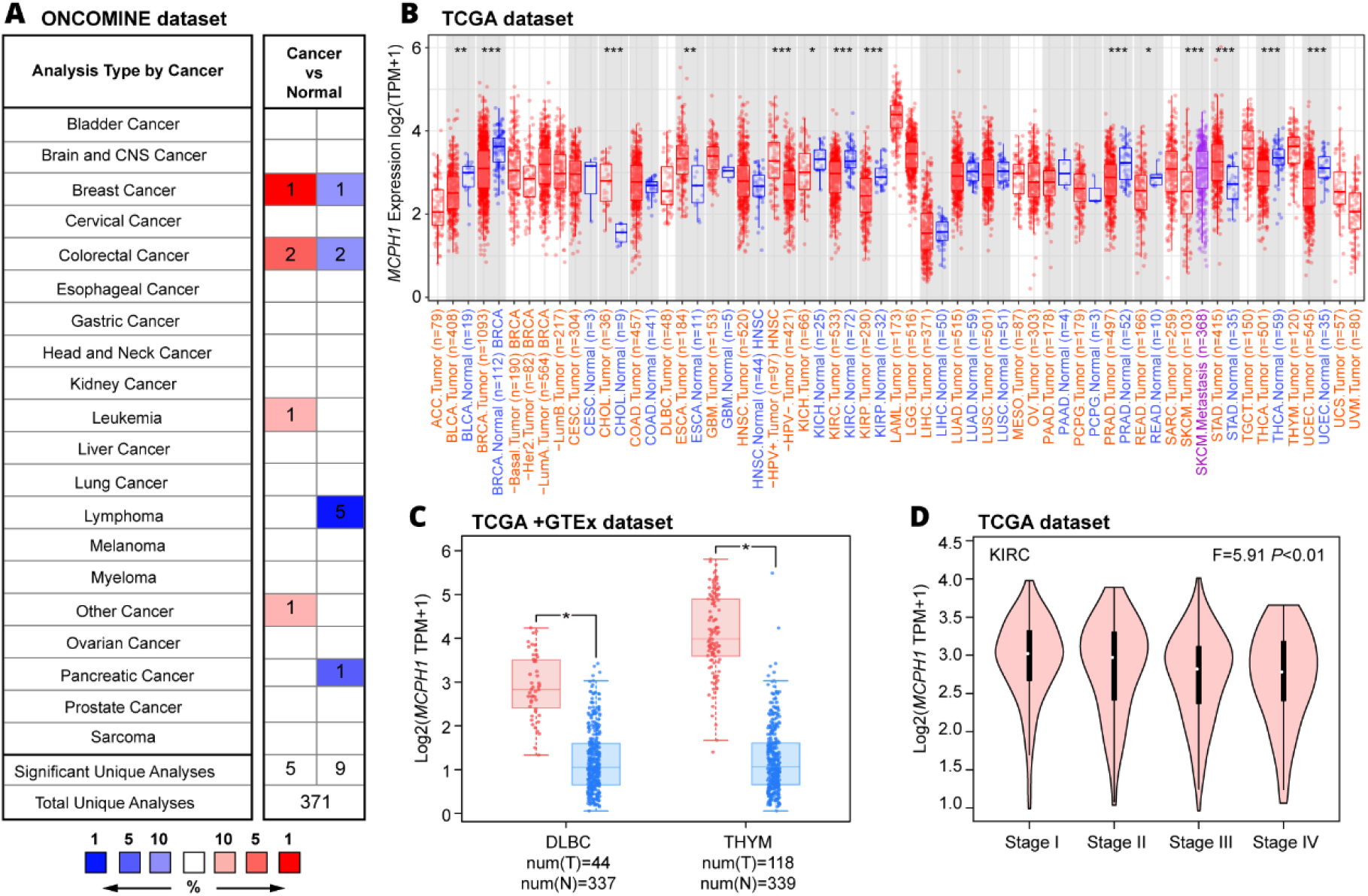
Expression level of microcephalin (*MCPH1*) gene in different tumors and pathological stages. (**A**) Increased or decreased expression of *MCPH1* in different cancer tissues, compared with normal tissues in ONCOMINE. Number in each cell is the amount of datasets. (**B**) MCPH1 expression levels in different cancer types from TCGA data in TIMER. \**P* < 0.05, \*\**P* < 0.01, \*\*\**P* < 0.001. (**C**) Box plot representation of *MCPH1* expression level comparison in DLBC and THYM (TCGA project relative to the corresponding normal tissues (GTEx database). \**P* < 0.05. (**D**) Based on the TCGA data, the expression levels of the MCPH1 gene were analyzed by the main pathological stages (stage I, stage II, stage III, and stage IV) of KIRC. Log2 (TPM + 1) was applied for log-scale.

For tumors lacking normal controls in TCGA, we analyzed these tumors using the GTEx database and showed that DLBC and THYM had elevated expression compared to normal tissues (**Fig. 1C**), while no significant differences were found in other tumors such as ACC, LAML, etc (**Fig. S4**).

We also explored the association between the expression of *MCPH1* and pathological stage by GEPIA2, which showed significant differences in KIRC (**Fig. 1D**) and no significant differences were found in other tumors (**Fig. S5**).

### Survival analysis

Relative expression of *MCPH1* in cancer cases was divided into the high and low expression to further investigate the correlation between abnormal *MCPH1* expression and prognosis of patients with different tumors. The results (**Fig. 2A**) demonstrate that *MCPH1* is highly expressed in LAML (*p*= 0.023), LIHC (Liver hepatocellular carcinoma *p*= 0.018), PAAD (*p*=0.049) and associated with poor prognosis of overall survival (OS). Disease-free survival (DFS) analysis (**Fig. 2B**) revealed a link between high *MCPH1* expression and poor prognosis in BLCA (*p*=0.012). Meanwhile, lower expression of the *MCPH1* gene was related to poor OS prognosis for KIRC (**Fig. 2A** *p*=3.2e-05) and DFS prognosis for KIRC (*p*= 0.016) and THCA (Thyroid carcinoma *p*=0.035) as shown in **Fig. 2B**.

**Fig. 2.**
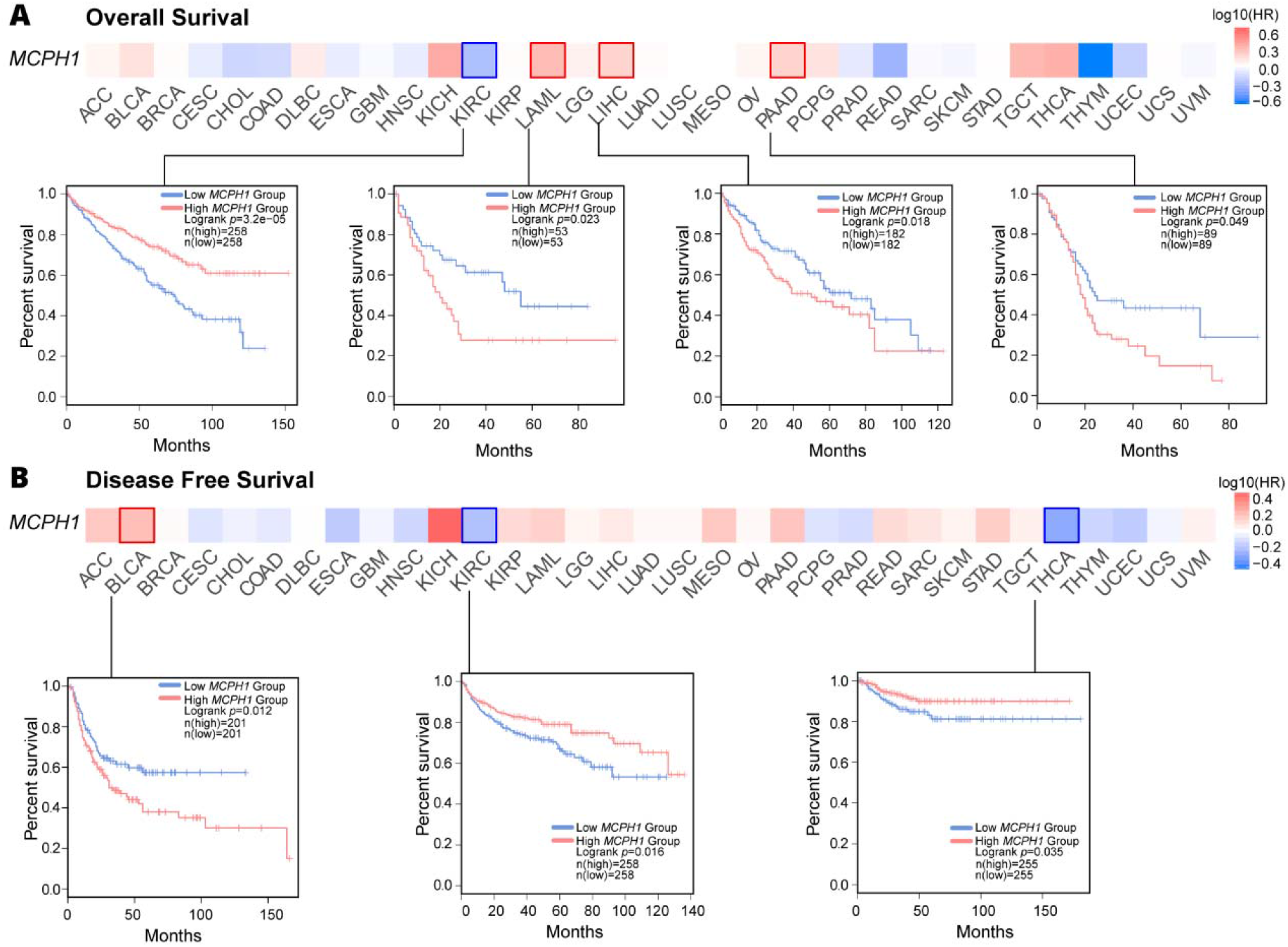
Correlation between *MCPH1* gene expression and survival prognosis of cancers in TCGA. GEPIA2 tool was used to perform overall survival (**A**) and disease-free survival (**B**) analyses of different tumors in TCGA by *MCPH1* gene expression. The survival map and Kaplan-Meier curves with positive results are given.

Moreover, Kaplan-Meier survival analysis of cancer cases was generated using Kaplan-Meier plotter tool. **Fig. S6** presented the relationship between lower expression of *MCPH1* and poor DMFS for breast cancer (**Fig. S6A)**, poor OS, FP (First progression) and PPS (Post-progression survival) prognosis for gastric cancer (**Fig. S6B)**, and poor OS, PFS (Relapse-free survival), PPS for ovarian cancer (**Fig. S6E)**.

### Genetic alteration analysis

TCGA cohort was used to explore different tumor samples and their genetic alteration status with *MCPH1*. The results revealed that uterine tumors, with “mutation” as the major type, have the highest frequency of *MCPH1* alteration (>8%). Liver cancer cases exhibited the highest incidence of “Deep Deletion” type of CNAs, with the frequency of ∼7% (**Fig. 3A**). **Fig. 3B** showed more detailed mutations and their locations within *MCPH1*. We did not find major types of genetic alteration and their location appeared to be somewhat sporadic. For example, a truncating mutation, T468Pfs*32 alteration, in the Microcephalin domain, was detected in 4 cases of STAD, 2 cases of UCEC, 1 case of COADREAD, and 1 case of CESE. We next explored the potential relationship between genetic alterations of *MCPH1* and the clinical survival prognosis of patients with different tumor types. The patients with genetic alteration of *MCPH1* do not show significant difference in DFS (*P*=0.060), PFS (*P*=0.078), OS (*P*=0.236), and disease specific survival (*P*=0.380), compared with patients without *MCPH1* alterations (**Fig. 3C**). Additionally, the expression of *MCPH1* was positively associated with TMB in COAD, LGG, and UCEC (*P* < 0.001), while negatively associated with TMB in THCA, THYM, BRCA, and PRAD cohorts (*P* < 0.001; **Fig. S7**). The expression of *MCPH1* is positively associated with MSI in UCEC, STAD, and READ cohorts (*P* < 0.001, **Fig. S8**). Taken together, *MCPH1* genetic alterations may be a potential driver of the above listed tumors.

**Fig. 3.**
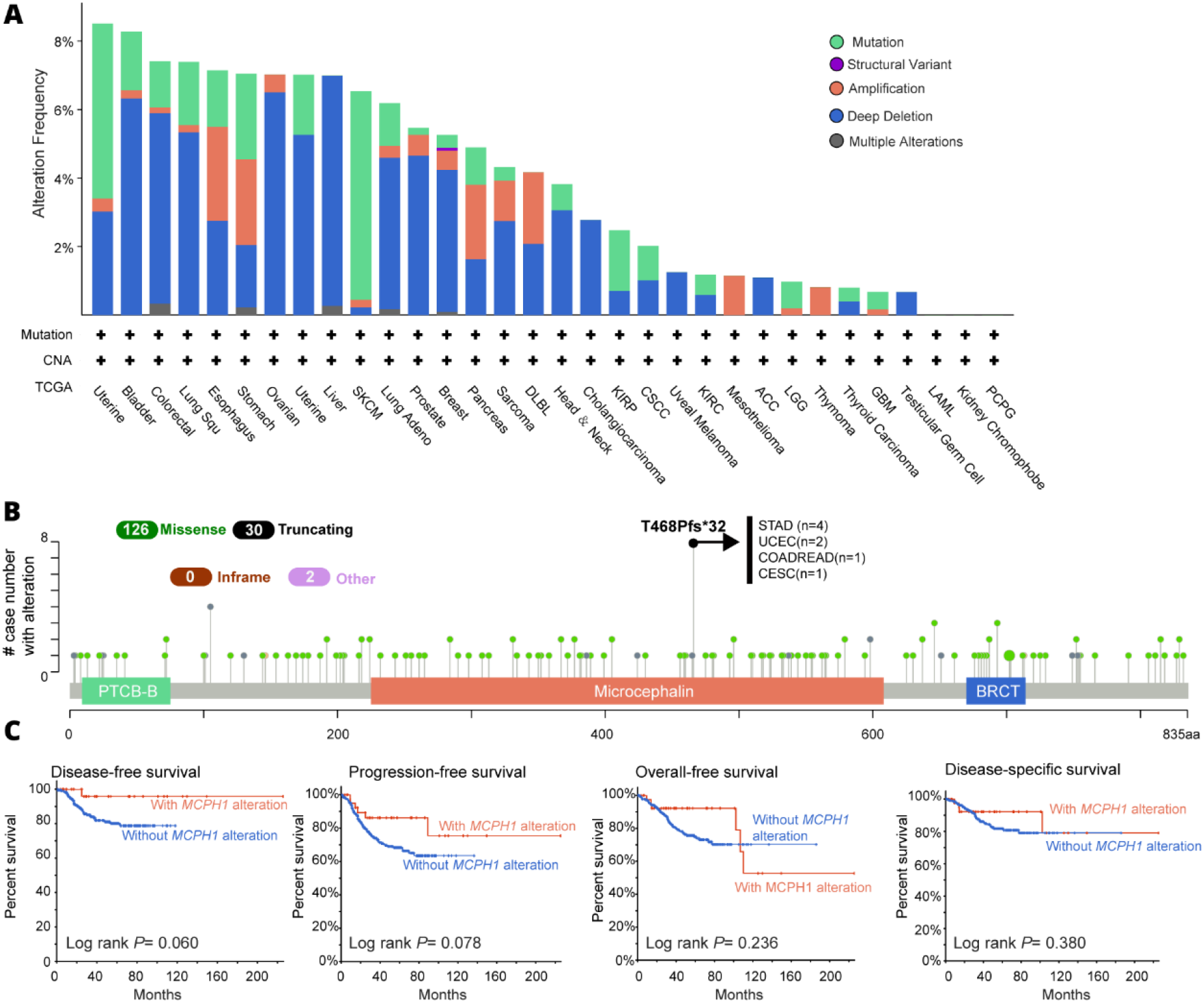
Mutation feature of *MCPH1* in different tumors of TCGA. We analyzed the mutation features of *MCPH1* for the TCGA tumors using the cBioPortal tool. The alteration frequency with mutation type (**A**) and mutation site (**B**) are displayed. We also analyzed the potential correlation between *MCPH1* alterations and without *MCPH1* alterations in disease-free, progression-free, overall-free, and disease-specific survival (**C**) using the cBioPortal tool.

### Immune microenvironment analysis

Next, we explored the connection between *MCPH1* expression and immune checkpoint genes expression using the expression data of more than forty immune checkpoint genes commonly found in various tumors types. The data of **Fig. 4** shows that the expression of *MCPH1* was positively correlated with the expression levels of immune checkpoint genes in different types of tumors, especially HNSC, LIHC, and PAAD. These results suggest that *MCPH1* may play a role in regulating the pattern of tumor immunity by modulating the expression levels of these immune checkpoint genes in some tumors.

**Fig. 4.**
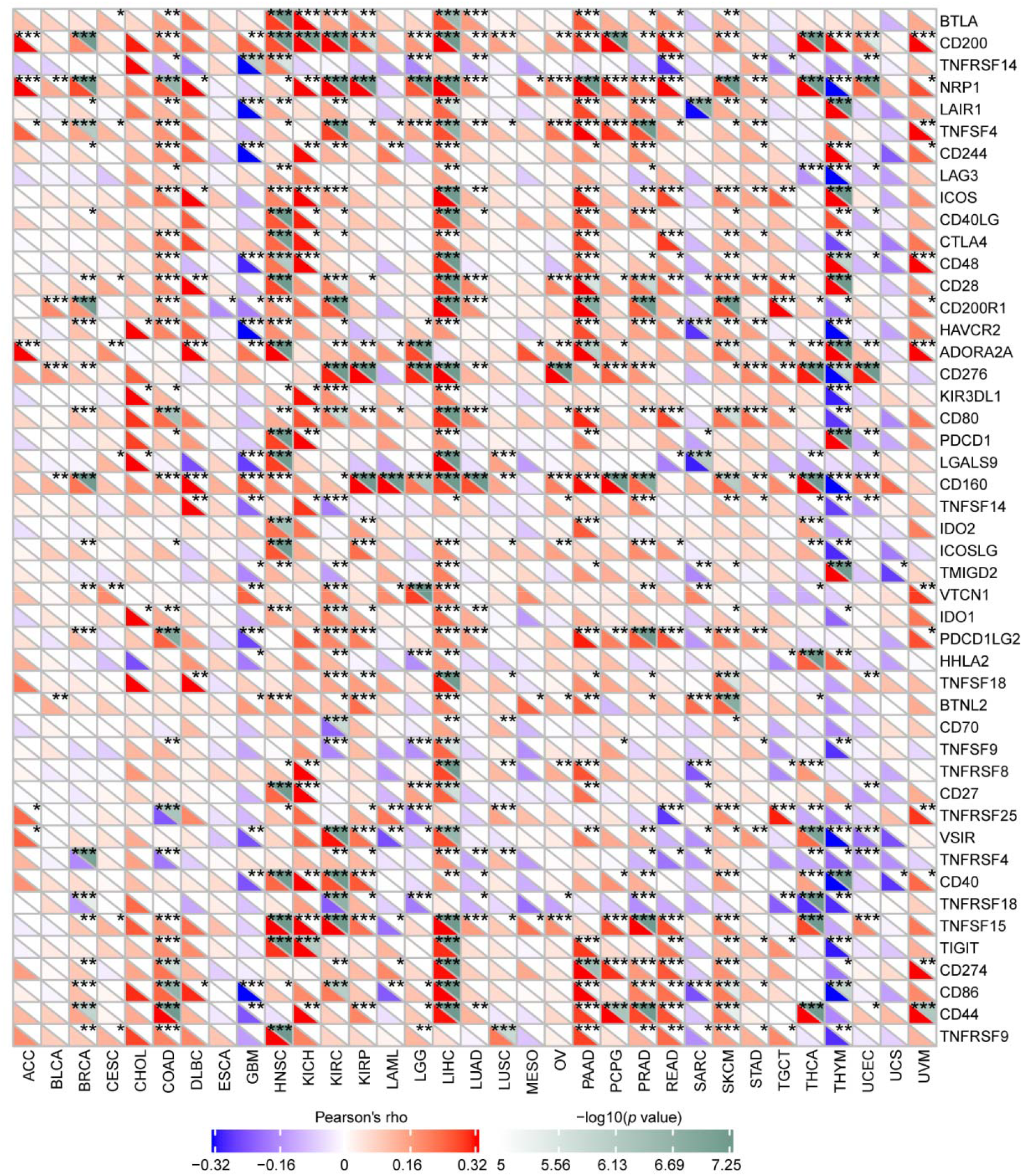
Correlations between *MCPH1* and immune checkpoints. The correlations between MCPH1 and confirmed immune checkpoints in multiple cancers (\**P* < 0.05, \*\**P* < 0.01, \*\*\**P* < 0.001).

To investigate whether *MCPH1* is involved in the pan-cancer immune infiltration process, we assessed the relationship between *MCPH1* expression and tumor purity. According to **Fig. 5**, *MCPH1* was most significantly associated with stromal scores in GBM, THYM, and SARC (Sarcoma). Meanwhile, the abundances of immune components in GBM, THCA, and SARC were significantly correlated with the expression level of *MCPH1*. Moreover, the relationships between *MCPH1* expression and tumor purity were significant in GBM, SARC, and THCA. To investigate the relationship between the expression of *MCPH1* and the number of antigens, we counted the number of nascent antigens for each tumor sample independently (**Fig. 6**). We found that the expression of *MCPH1* was positively correlated with the number of neoantigens in STAD (R=0.169, *P*=0.009) and LGG (R=0.216, *P*=0.002).

**Fig. 5.**
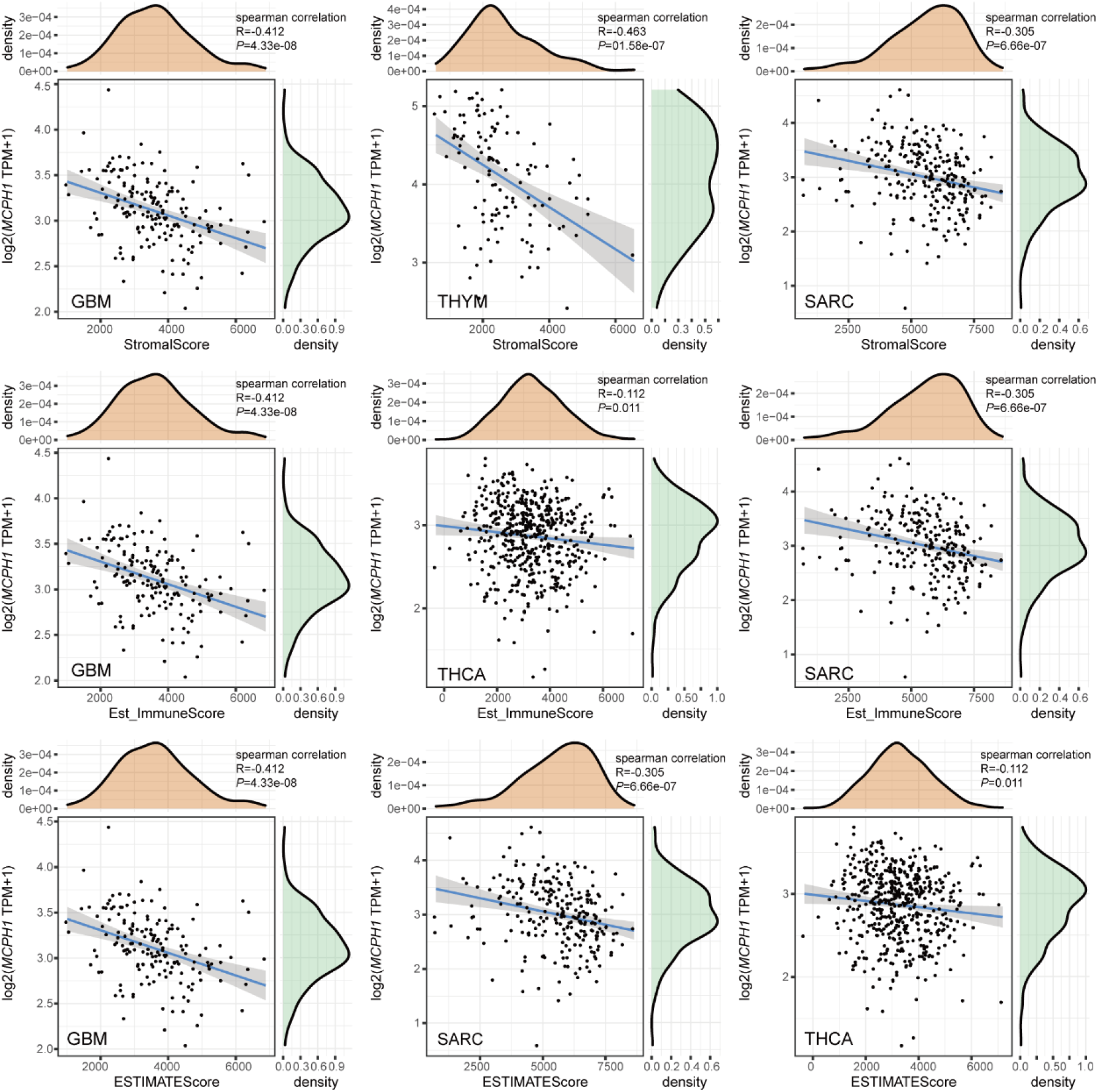
Correlations between *MCPH1* and tumor purity. Top three scatter plots of correlation between *MCPH1* and stromal score, immune score, ESTIMATE score in various cancers.

**Fig. 6.**
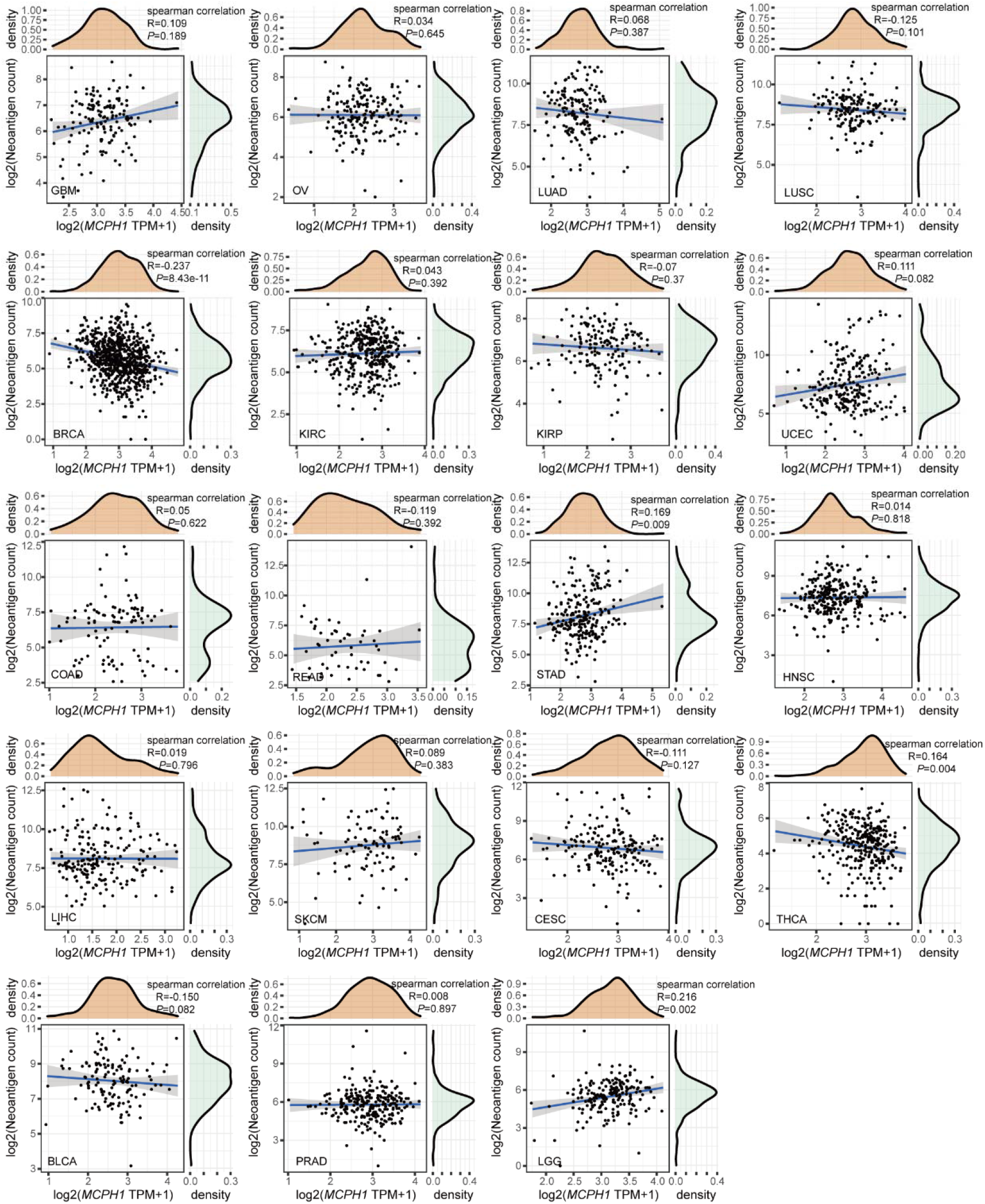
Correlation analysis between *MCPH1* expression and immune neoantigens.

In addition, we displayed the relationship between *MCPH1* and various immune infiltrates in human cancers using TIMER2.0 (**Fig. 7**). Overall, we found a positive correlation between *MCPH1* and the level of immune infiltrating of various infiltrating cells, such as Monocyte, Neutrophil, Mast, Endo, Common lymphoid progenitor, and MDSC. However, the expression of *MCPH1* was negatively correlated with NKT. Taken together, our results indicate that *MCPH1* may be highly involved in the process of immune infiltration and formation of diverse components in the above tumors.

**Fig. 7.**
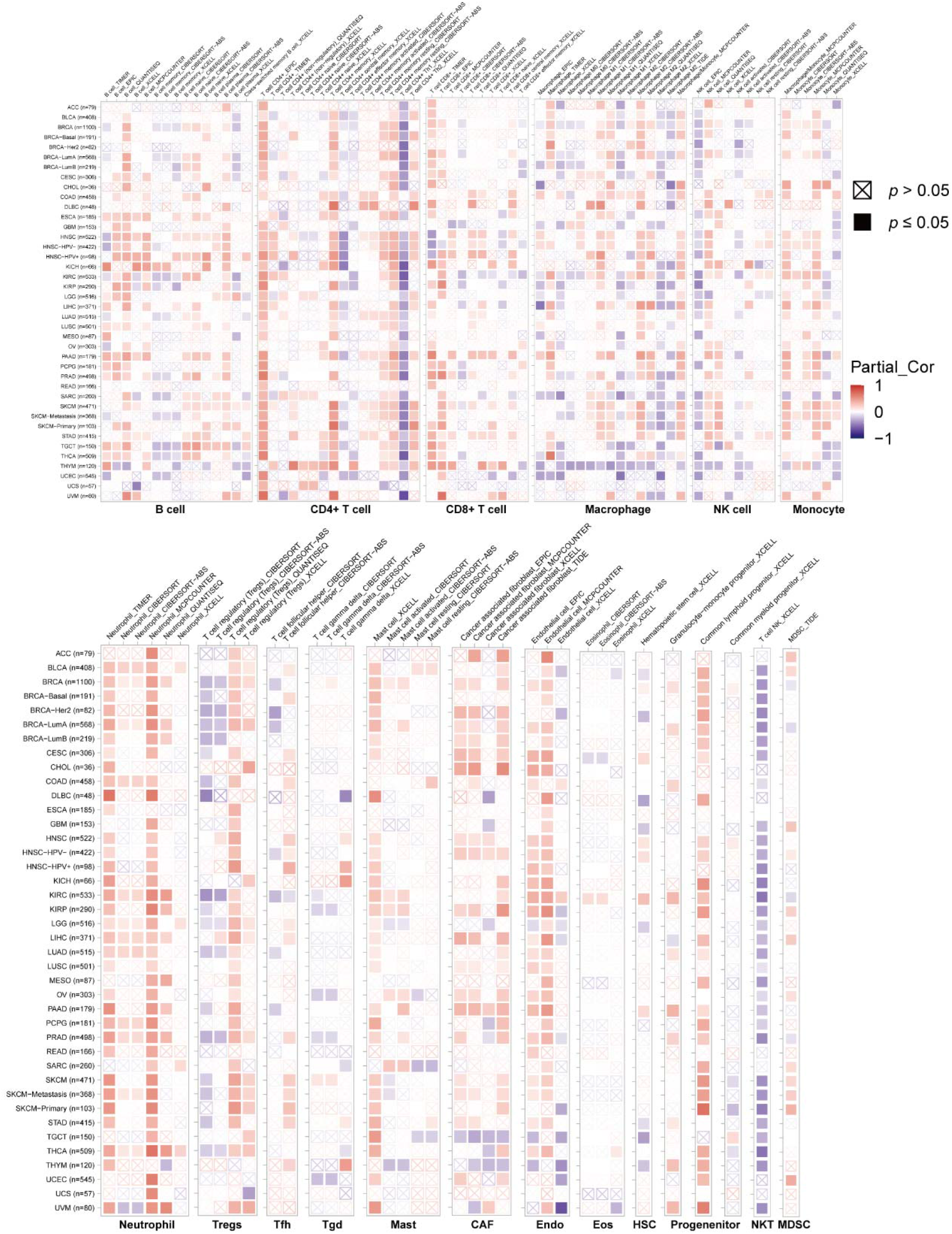
The correlations of *MCPH1* expression and immune infiltration in various cancers.

### Enrichment analysis of *MCPH1*-related partners

To investigate the potential molecular mechanism of the *MCPH1* gene in tumor development, we selected the MCPH1-interacting proteins and the *MCPH1* expression correlated genes for a series of pathway enrichment analyses. The data showed the interaction network of 40 MCPH1-interacting proteins (**Fig. 8A**). Then GEPIA2 tool was used to obtain the top 100 *MCPH1* expression correlated genes. The expression of *MCPH1* was positively correlated with that of *XPO7* (R=0.69), *PPP2R2A* (R=0.52), *CTCF* (R=0.62), *WRN* (R=0.59) and *CNOT7* (R=0.58) genes (all *P*<0.001) (**Fig. 8B**). The heatmap data displayed a strong positive association of *MCPH1* with all five of these genes in most cancer types (**Fig. 8C**).

**Fig. 8.**
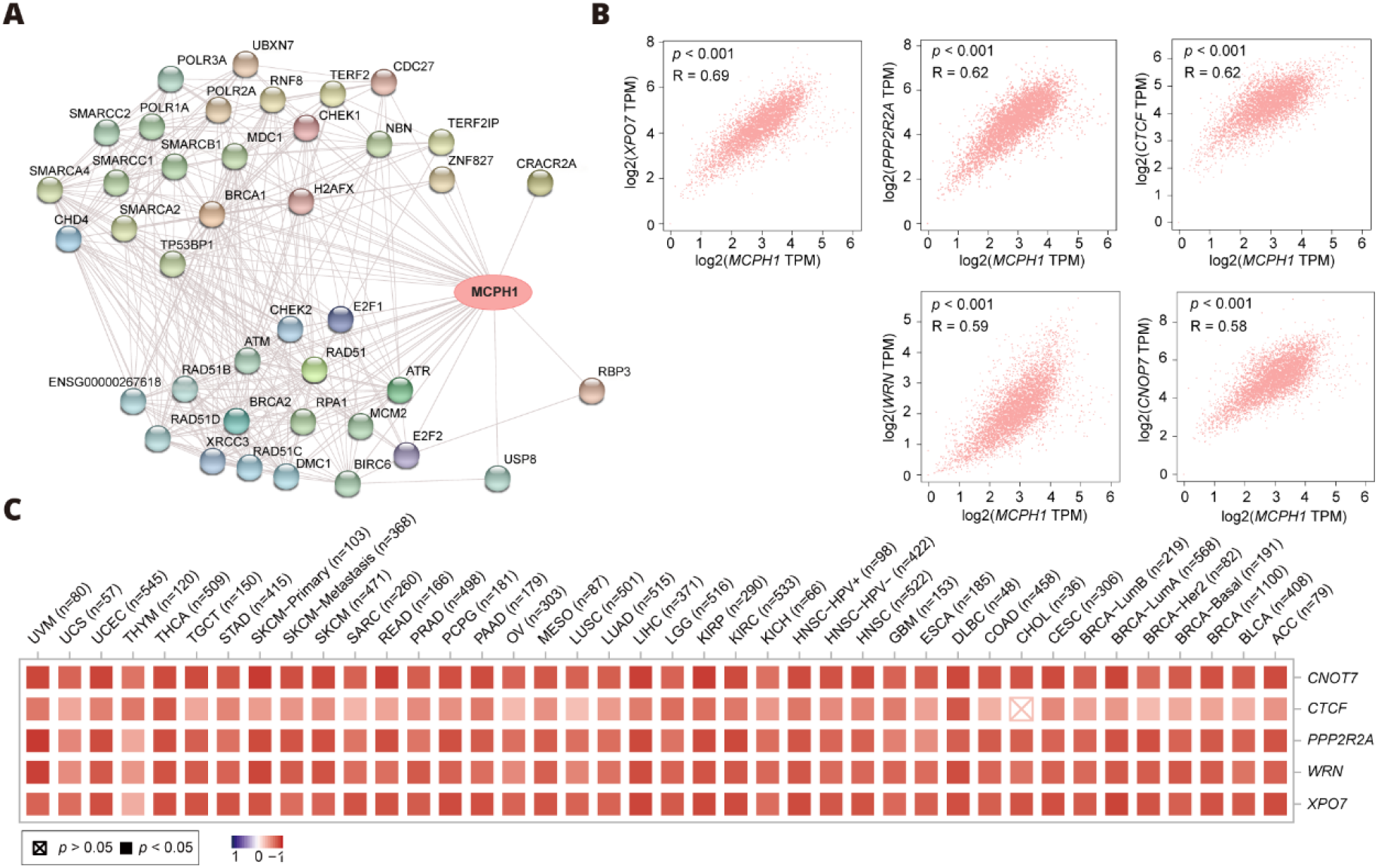
MCPH1-related gene enrichment analysis. (**A**) STRING protein network map of experimentally determined MCPH1-binding proteins. Colored nodes indicate the individual proteins identified. (**B**) Expression correlation between *MCPH1* and representative genes (*XPO7, PPP2R2A, CTCF, WRN* and *CNOT7*) of the top *MCPH1*-correlated genes in TCGA projects as determined by GEPIA2. (**C**) Heatmap representation of the expression correlation data between *MCPH1* and *XPO7, PPP2R2A, CTCF, WRN* and *CNOT7* in the TCGA tumors.

We integrated the two datasets to perform GO and KEGG enrichment analyses. GO enrichment suggests that most of these gene are mightily correlated with the pathways or cellular biology of DNA metabolism, such as helicase activity, single-stranded DNA binding, DNA helicase activity, and others (**Fig. S9A**). KEGG analysis suggests that “cell cycle” and “Homologous recombination” might be involved in the effect of MCPH1 on tumor pathogenesis (**Fig. S9B**).

## Discussion

Several reports have demonstrated that the multifunctional MCPH1 protein is involved in a variety of cellular biological processes across different mammalian species and cell lines, including DNA repair^24,25^, cell cycle^26,27^, and tumorigenesis^6,9-12,26^. In this paper, “HomoloGene” and phylogenetic tree analysis revealed that the conservation of the MCPH1 protein structure across different species, implying that similar mechanisms may exist for the normal physiological role of *MCPH1*. However, whether *MCPH1* contributes to the development of many cancers via shared molecular processes remains unknown. Hitherto, we found there were no relevant studies on pan-cancer analysis of *MCPH1* from the standpoint of total tumors. Therefore, we performed the first comprehensive analysis of the *MCPH1* gene in various tumors based on the data of TCGA, OMCOMINE and GEO databases, as well as molecular characterization of gene expression and immune infiltration.

The OMCOMINE and TIMER2.0 database analysis revealed a significant difference in the expression of the *MCPH1* gene between tumors and normal tissues. OMCOMINE data showed that *MCPH1* levels were decreased in breast, colorectal and pancreatic cancers, and lymphoma, which is in good agreement with previous cognition of *MCPH1* as a cancer suppressor gene. However, we found *MCPH1* level is surprisingly elevated in breast cancer, colorectal cancers, and leukemia compared to normal tissues. TCGA data exhibited that the expression level of *MCPH1* was higher in CHOL, ESCA, and STAD than in the corresponding control tissues, while lower expression in BLCA, BRCA, KICH, KIRC, KIRP, PRAD, THCA, UCEC, and READ. These results indicated that the abnormal expression levels of *MCPH1* including both elevation and descent in different tumor types might reflect distinct potential functions and mechanisms. We further revealed that for tumor patients with high *MCPH1* expression, such as LAML, LIHC, and PAAD, over-expression of *MCPH1* usually predicted poor OS. Taken together, our data imply that *MCPH1* might be a potential biomarker for predicting the prognosis of tumor patients.

It is worth mentioning that there were some interesting findings on oncogenic role of *MCPH1* in our analysis which are previously underexplored. For instance, the role of *MCPH1* in gastric cancer was rarely reported in prior studies. Our analysis revealed that low expression of *MCPH1* was correlated with poor prognosis of OS, FP, and PPS in gastric cancer, indicating that *MCPH1* might be a novel potential clinical biomarker for predicting the overall survival of gastric cancer. However, additional basic and clinical experiments are required to corroborate this hypothesis.

For kidney renal clear cell carcinoma (KIRC), our analysis demonstrated a correlation between low expression of *MCPH1* and poor OS prognosis (*p*<0.001), poor DFS (*P*=0.017), late clinical staging (*P*<0.01). This finding is consistent with a prior publication indicating that *MCPH1* is a novel molecular marker for human kidney cancer^28^.

Additionally, we discovered that the expression of *MCPH1* is strongly related with the immune microenvironment, which has rarely been reported in the literature. The expression of *MCPH1* is positively associated with immune infiltrates, especially Monocyte, Neutrophil, Mast, Endo, Common lymphoid progenitor, and MDSC in various cancers, while it is negatively correlated with NKT. Besides, we discovered that *MCPH1* expression is positively correlated with the number of neoantigens in STAD and LGG. We assumed that *MCPH1* would mediate similar immune activities in various tumors based on our results. However, further work should be required to determine whether *MCPH1* performs such functions. Despite the significance of this correlation between the expression of *MCPH1* and MSI or TMB, the correlation coefficients between *MCPH1* and TMB, as well as MSI, were below 0.6 in almost all cancers, indicating that *MCPH1* was unlikely to influence tumorigenesis via genetic modifications. Further results of enrichment analyses revealed that *MCPH1* may be indirectly involved in tumor formation during the DNA damage induction response. Nevertheless, this conjecture needs further experimental verification. On the whole, our new findings may provide a new therapeutic strategy for malignancies mediated by *MCPH1*.

## Conclusion

To conclude, our pan-cancer analysis of *MCPH1* indicated statistical correlations of abnormal *MCPH1* expression with clinical prognosis, immune microenvironment, tumor mutational burden or microsatellite instability across different tumors, which contributes to understanding the potential molecular mechanisms of *MCPH1* in tumorigenesis. Moreover, our study indicated that *MCPH1* may be a valuable biomarker candidate for cancer diagnosis, prognosis, therapy design, and follow-up, especially in LAML, PAAD and gastric cancer.

## Data Availability

All data produced in the present work are contained in the manuscript

https://www.oncomine.org/

http://timer.cistrome.org/

http://gepia2.cancer-pku.cn/#analysis

https://www.cbioportal.org/

http://vip.sangerbox.com/

http://string-db.org/

http://genome.ucsc.edu/

http://www.proteinatlas.org/

https://www.aclbi.com/

http://kmplot.com/analysis/

## Data availability

The datasets generated and analyzed during the current study are available from the corresponding author on reasonable request.

## Acknowledgments

The authors gratefully acknowledge the Gene Expression Omnibus (GEO) database, and the Cancer Genome Atlas (TCGA) database, which made the data available. We are very greatful to associate professor Chengqiang Feng for his help in revising the manuscript.

## Authors’ contributions

This research was conducted in collaboration with all authors. ZW and ML performed the data analysis and visualization, and ZW drafted the manuscript. WD, JH, WK, ZL, and YC critically reviewed and revised this manuscript. All authors read and approved the final manuscript.

## Additional information

Supplementary Information: The online version contains supplementary material available at https://doi.org/10.1038/XXX.

Correspondence and requests for materials should be addressed to J.H. or W.D.

### Abbreviations

ACC: Adrenocortical carcinoma
BLCA: Bladder urothelial carcinoma
BRCA: Breast invasive carcinoma
BRCT: BRCA1 C-terminus
CESC: Cervical squamous cell carcinoma
CHOL: Cholangiocarcinoma
CNAs: Copy number alterations
COAD: Colon adenocarcinoma
COR: correlation
DFS: Disease-free survival
DLBC: Diffuse large B cell lymphoma
DMFS: distant metastasis-free survival
DSS: disease-specific survival
ESCA: Esophageal carcinoma
FC: Fold change
FP: First progression
GBM: Glioblastoma multiforme
GEO: Gene Expression Omnibus
GEPIA: Gene Expression Profiling Interactive Analysis
GO: Gene ontology
GTEx: Genotype-Tissue Expression
HNSC: Head and neck squamous cell carcinoma
HPA: Human Protein Atlas
KEGG: Kyoto encyclopedia of genes and genomes
KICH: Kidney chromophobe
KIRC: Kidney renal clear cell carcinoma
KIRP: Kidney renal papillary cell carcinoma
LAML: Acute myeloid leukemia
LGG: Brain lower grade glioma
LIHC: Liver hepatocellular carcinoma
LUAD: Lung adenocarcinoma
LUSC: Lung squamous cell carcinoma
MESO: Mesothelioma
MSI: microsatellite instability
NX: Normalized expression
OS: Overall survival
OV: Ovarian serous cystadenocarcinoma
PAAD: Pancreatic adenocarcinoma
PCPG: Pheochromocytoma and paraganglioma
PFS: progress-free survival
PPS: post-progression survival
PRAD: Prostate adenocarcinoma
READ: Rectum adenocarcinoma
RFS: relapse-free survival
SARC: Sarcoma
SKCM: Skin cutaneous melanoma
STAD: Stomach adenocarcinoma
TGCT: Testicular germ cell tumors
TPM: Transcripts per million
TIMER: tumor immune estimation resource
THCA: Thyroid carcinoma
THYM: Thymoma
UCEC: Uterine corpus endometrial carcinoma
UCS: Uterine carcinosarcoma
UVM: Uveal melanoma

## References

1. Liu, Y. et al., Pan-cancer analysis on the role of PIK3R1 and PIK3R2 in human tumors. Sci Rep-Uk 12 5924 (2022).

2. de Reyniès, A. et al., Large-scale pan-cancer analysis reveals broad prognostic association between TGF-β ligands, not Hedgehog, and GLI1/2 expression in tumors. Sci Rep-Uk 10 14491 (2020).

3. Wen, J., Mao, X., Cheng, Q., Liu, Z. & Liu, F., A pan-cancer analysis revealing the role of TIGIT in tumor microenvironment. Sci Rep-Uk 11 22502 (2021).

4. Chen, D. et al., Pan-cancer analysis of the prognostic and immunological role of PSMB8. Sci Rep-Uk 11 20492 (2021).

5. Brüning-Richardson, A. et al., ASPM and microcephalin expression in epithelial ovarian cancer correlates with tumour grade and survival. Brit J Cancer 104 1602 (2011).

6. Rai, R. et al., BRIT1 regulates early DNA damage response, chromosomal integrity, and cancer. Cancer Cell 10 145 (2006).

7. Hagemann, C. et al., Expression analysis of the autosomal recessive primary microcephaly genes MCPH1 (microcephalin) and MCPH5 (ASPM, abnormal spindle-like, microcephaly associated) in human malignant gliomas. Oncol Rep 20 301 (2008).

8. Liu, X. et al., The N-terminal BRCT domain determines MCPH1 function in brain development and fertility. Cell Death Dis 12 143 (2021).

9. Richardson, J. et al., Microcephalin is a new novel prognostic indicator in breast cancer associated with BRCA1 inactivation. Breast Cancer Res Tr 127 639 (2011).

10. Jo, Y. H. et al., MCPH1 protein expression and polymorphisms are associated with risk of breast cancer. Gene 517 184 (2013).

11. Liang, Y. et al., Mcph1/Brit1 deficiency promotes genomic instability and tumor formation in a mouse model. Oncogene 34 4368 (2015).

12. Wu, X., Liu, W., Liu, X., Ai, Q. & Yu, J., Overexpression of MCPH1 inhibits the migration and invasion of lung cancer cells. Oncotargets Ther Volume 11 3111 (2018).

13. Clough, E. & Barrett, T., (Springer New York, kNew York, NY, 2016), Vol. 1418, pp. 93.

14. Blum, A., Wang, P. & Zenklusen, J. C., SnapShot: TCGA-Analyzed Tumors. Cell 173 530 (2018).

15. Tomczak, K., Czerwińska, P. & Wiznerowicz, M., Review The Cancer Genome Atlas (TCGA): an immeasurable source of knowledge. Współczesna Onkologia 1A 68 (2015).

16. Rhodes, D. R. et al., ONCOMINE: A Cancer Microarray Database and Integrated Data-Mining Platform. Neoplasia (New York, N.Y.) 6 1 (2004).

17. Tang, Z., Kang, B., Li, C., Chen, T. & Zhang, Z., GEPIA2: an enhanced web server for large-scale expression profiling and interactive analysis. Nucleic Acids Res 47 W556 (2019).

18. Gao, J. et al., Integrative Analysis of Complex Cancer Genomics and Clinical Profiles Using the cBioPortal. Sci Signal 6 l1 (2013).

19. Cerami, E. et al., The cBio Cancer Genomics Portal: An Open Platform for Exploring Multidimensional Cancer Genomics Data: Figure 1. Cancer Discov 2 401 (2012).

20. Ye, W., Luo, C., Liu, F., Liu, Z. & Chen, F., CD96 Correlates With Immune Infiltration and Impacts Patient Prognosis: A Pan-Cancer Analysis. Front Oncol 11 (2021).

21. Li, T. et al., TIMER: A Web Server for Comprehensive Analysis of Tumor-Infiltrating Immune Cells. Cancer Res 77 e108 (2017).

22. Li, T. et al., TIMER2.0 for analysis of tumor-infiltrating immune cells. Nucleic Acids Res 48 W509 (2020).

23. Szklarczyk, D. et al., STRING v11: protein–protein association networks with increased coverage, supporting functional discovery in genome-wide experimental datasets. Nucleic Acids Res 47 D607 (2019).

24. Liang, Y. et al., BRIT1/MCPH1 Is Essential for Mitotic and Meiotic Recombination DNA Repair and Maintaining Genomic Stability in Mice. Plos Genet 6 e1000826 (2010).

25. Yang, S. Z., Lin, F. T. & Lin, W. C., MCPH1/BRIT1 cooperates with E2F1 in the activation of checkpoint, DNA repair and apoptosis. Embo Rep 9 907 (2008).

26. Zhou, L. et al., Overexpression of MCPH1 inhibits uncontrolled cell growth by promoting cell apoptosis and arresting the cell cycle in S and G2/M phase in lung cancer cells. Oncol Lett 11 365 (2016).

27. Mai, L. et al., The overexpression of MCPH1 inhibits cell growth through regulating cell cycle-related proteins and activating cytochrome c-caspase 3 signaling in cervical cancer. Mol Cell Biochem 392 95 (2014).

28. Wang, N. et al., Primary microcephaly gene MCPH1 shows a novel molecular biomarker of human renal carcinoma and is regulated by miR-27a. Int J Clin Exp Patho 7 4895 (2014).

