## Additional information for "The Oncogenic Role of Human Microcephalin Gene Revealed by Pan-Cancer Analysis"

**Supplementary materials and methods**

1. **Gene mapping and protein structure analysis**

We obtained *MCPH1* genome location information using UCSC human genome browser (http://genome.ucsc.edu/). NCBI (National Center for Biotechnology Information) was used to conduct an analysis of conserved functional domain and the phylogenetic tree of *MCPH1* in different species.

2. **Gene expression analysis of HPA**

HPA (Human Protein Atlas) database (http://www.proteinatlas.org/) was used to explore the expression level of the *MCPH1* under physiological conditions in different cell- and tissue types. “Low specificity” was defined by “NX (Normalized expression) ≥1 in at least one tissue/region/cell type but not elevated in any tissue/region/cell type”.

3. **Survival prognosis analysis of Kaplan-Meier plotter**

We pooled the different GEO datasets for a string of analyses of OS, DMFS (distant metastasis-free survival), RFS (relapse-free survival), PPS (post-progression survival), FP (first progression), DSS (disease-specific survival), and PFS (progress-free survival) using Kaplan-Meier plotter (http://kmplot.com/analysis/).

4. **Correlation of MCPH1 and TMB/MSI**

The potential correlation between *MCPH1* expression and TMB (tumor mutational burden) or MSI (microsatellite instability) in TCGA tumors was analyzed using the web tool: “https://www.aclbi.com/”.

**Supplementary Figures**

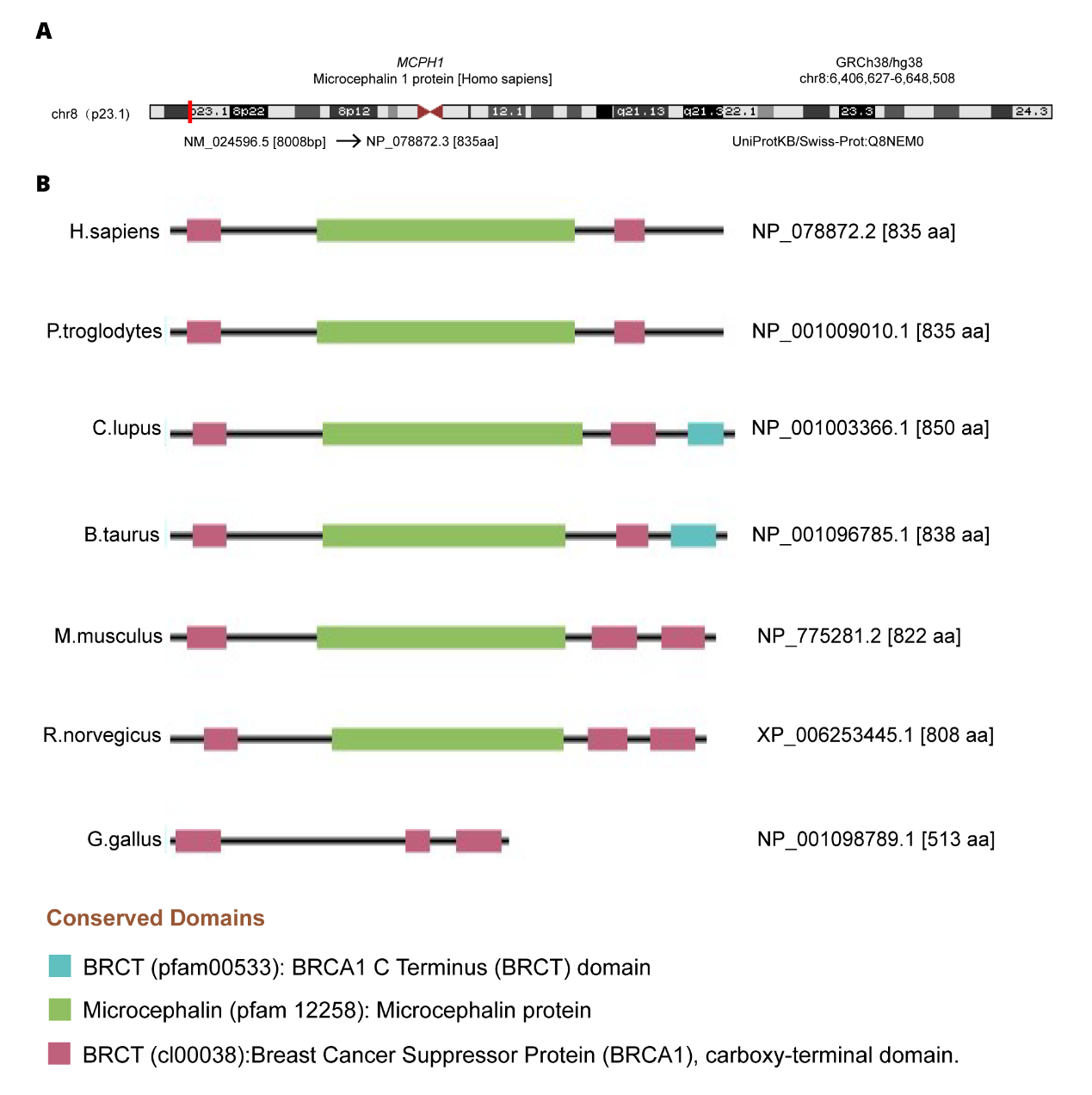

**Fig. S1** Structural characteristics of *MCPH1* in different species. (**A**) Genomic location of human *MCPH1*; (**B**) Conserved domains of MCPH1 protein among different species.

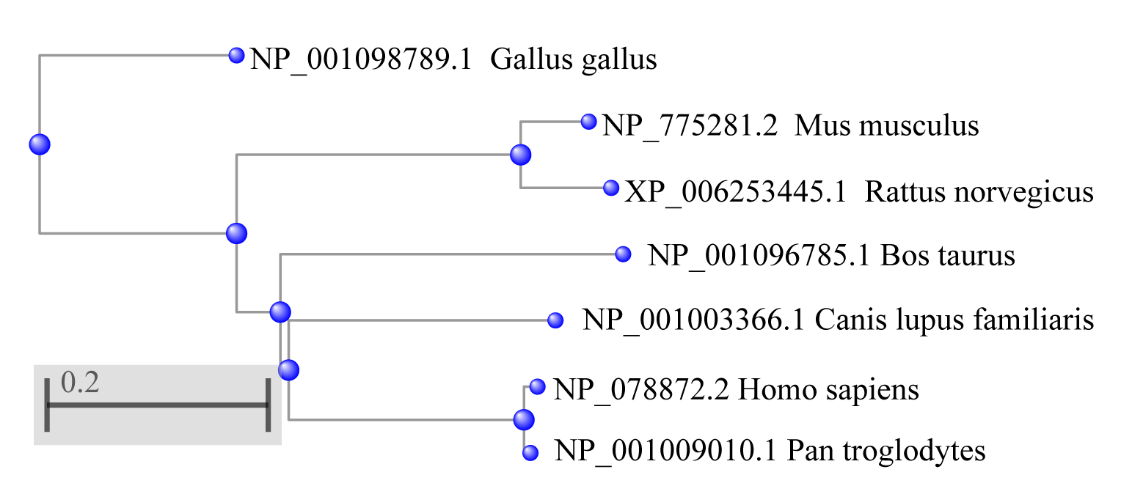

**Fig. S2**: Phylogenetic tree of MCPH1. We used a constraint-based multiple alignment tool of NCBI to obtain the phylogenetic tree of MCPH1 in different species.

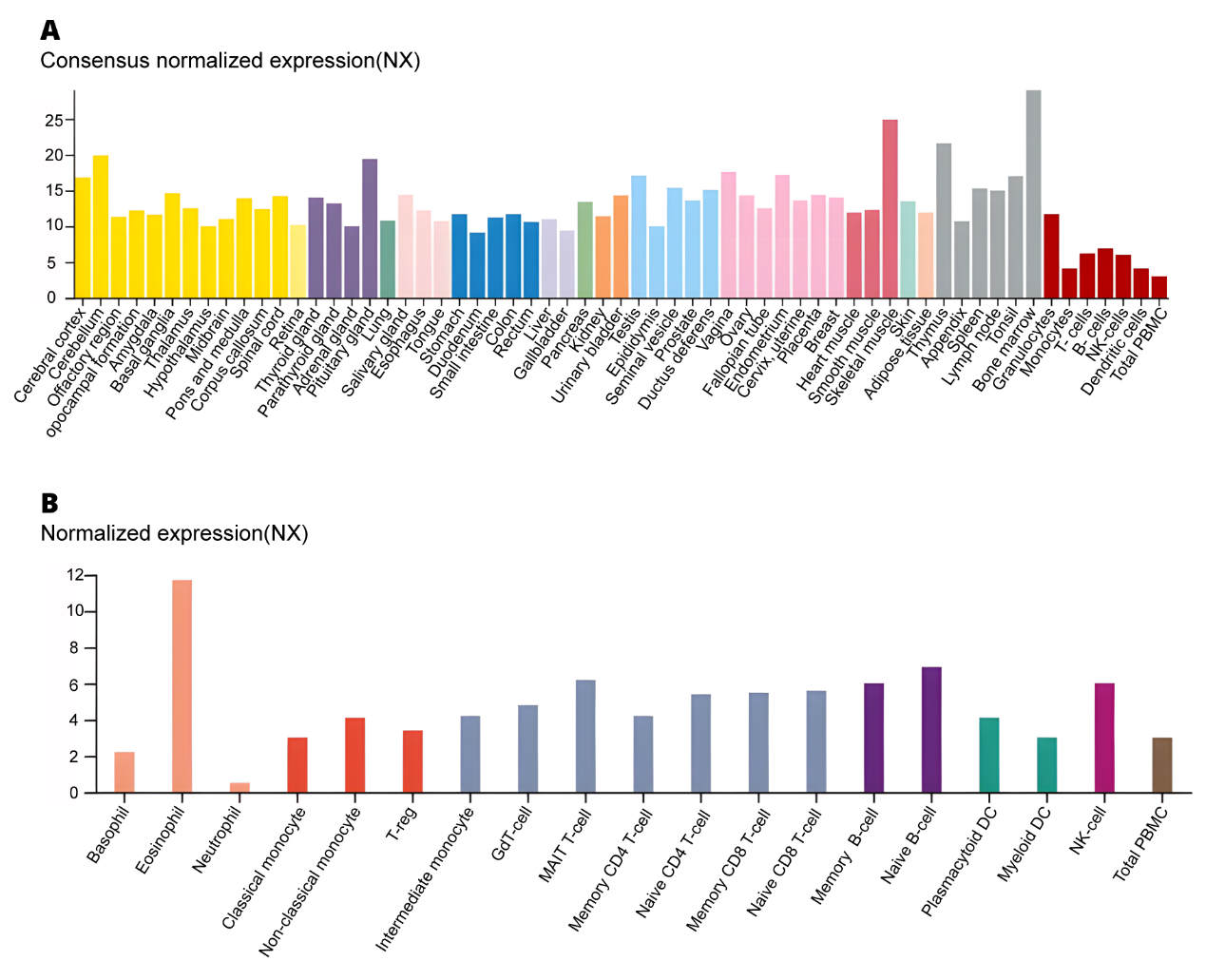

**Fig. S3**: Expression level of *MCPH1* in different cells and tissues under normal physiological conditions. (**A**) Expression of *MCPH1* gene in different tissues based on HPA, GTEx and FANTOM5 database; (**B**) Expression of *MCPH1* gene in different blood cell types based on the HPA, Monaco and Schmiedel datasets.

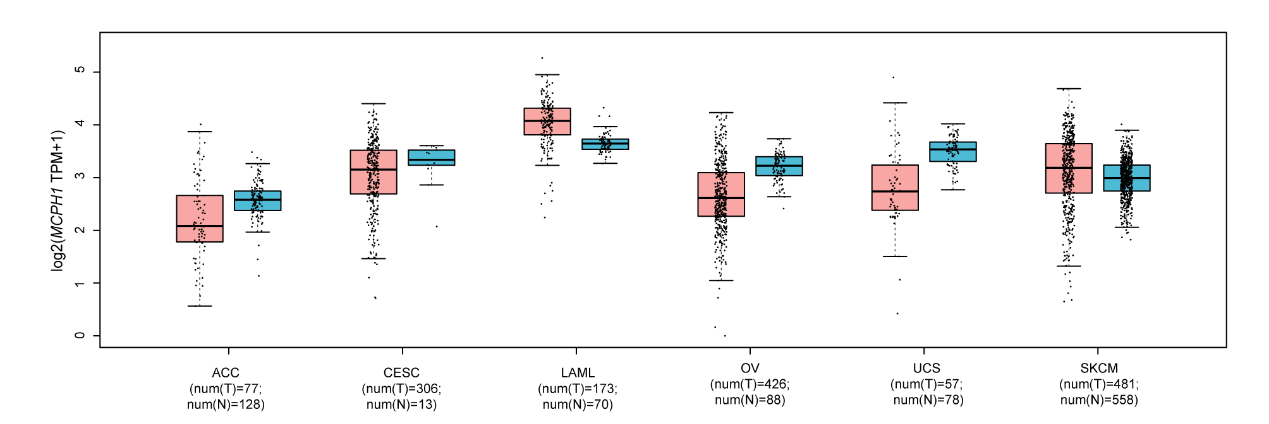

**Fig. S4**: Comparison of *MCPH1* expression levels using GTEx database. The expression statuses of the *MCPH1* gene in tumors without available appropriate control in TCGA, such as ACC, CESC, LAML, OV, UCS, SKCM, were compared to normal tissues based on the GTEx databases.

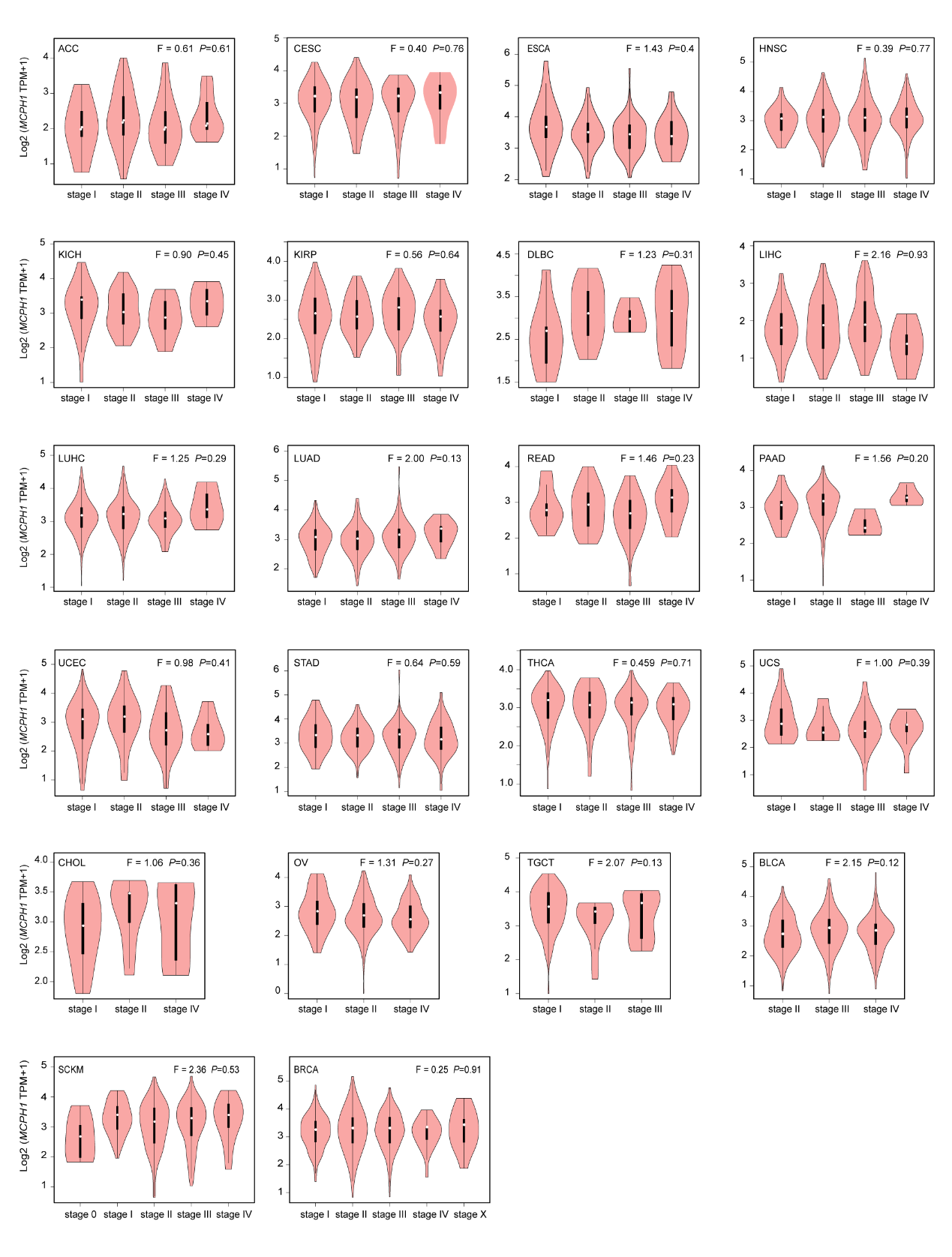

**Fig. S5**: Relationship between *MCPH1* expression level and pathological stages of TCGA tumors. Pathological stage plot derived for *MCPH1* gene expression data in GEPIA2 for ACC, BLCA, BRCA, CESC, CHOL, DLBC, ESCA, HNSC, KICH, LIHC, LUAD, LUHC, OV, PAAD, READ, SCKM, STAD, TGCT, THCA, UCEC and UCS.

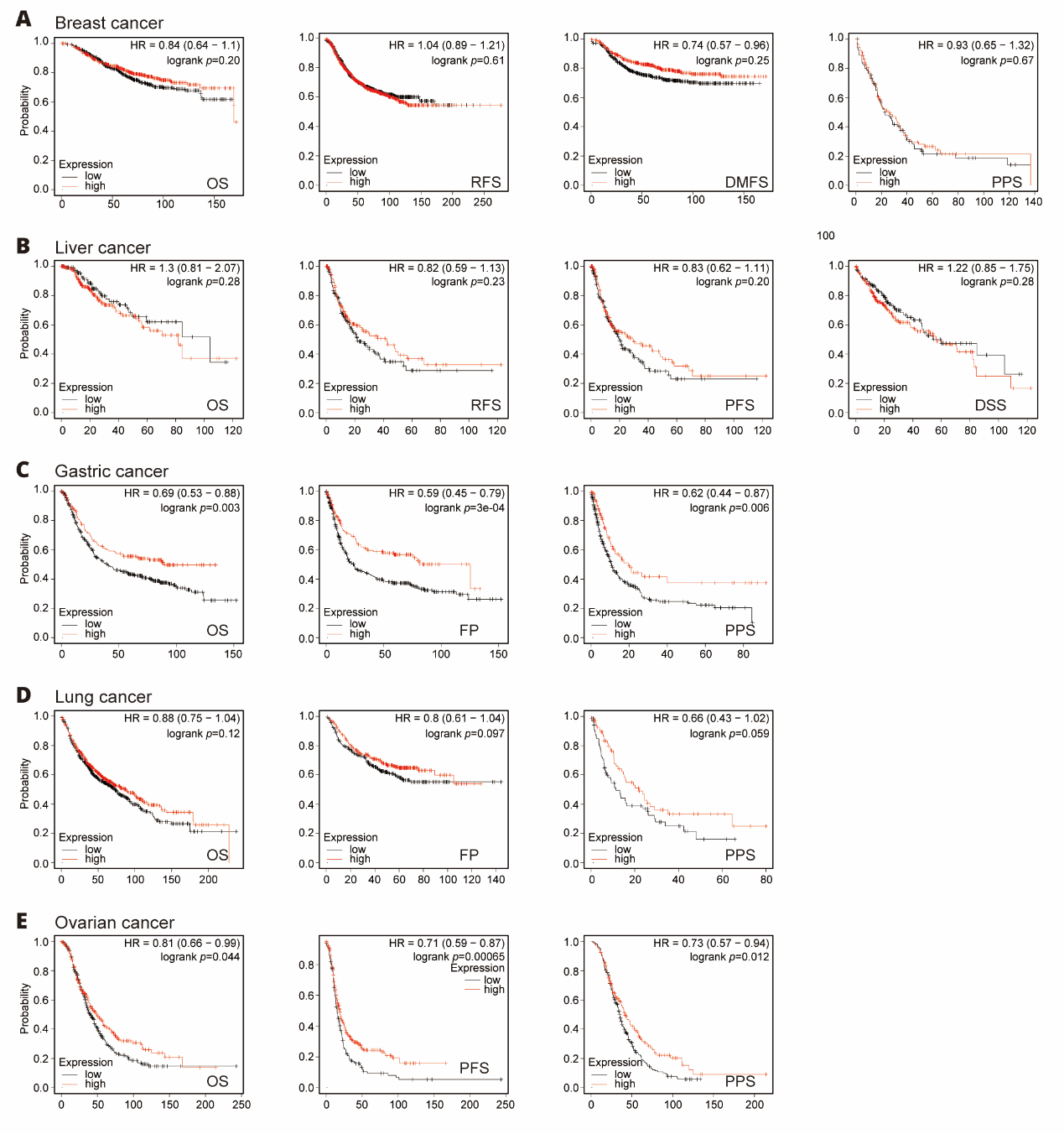

**Fig. S6**: Correlation between *MCPH1* gene expression and prognosis of cancers using the Kaplan-Meier plotter. We used the Kaplan-Meier plotter to perform a series of survival analyses, including OS, DMFS, RFS, PFS, PPS, FP, and DSS, via the expression level of the *MCPH1* gene in breast cancer (**A**), liver cancer (**B**), gastric cancer (**C**), lung cancer (**D**), and ovarian cancer (**E)** cases.

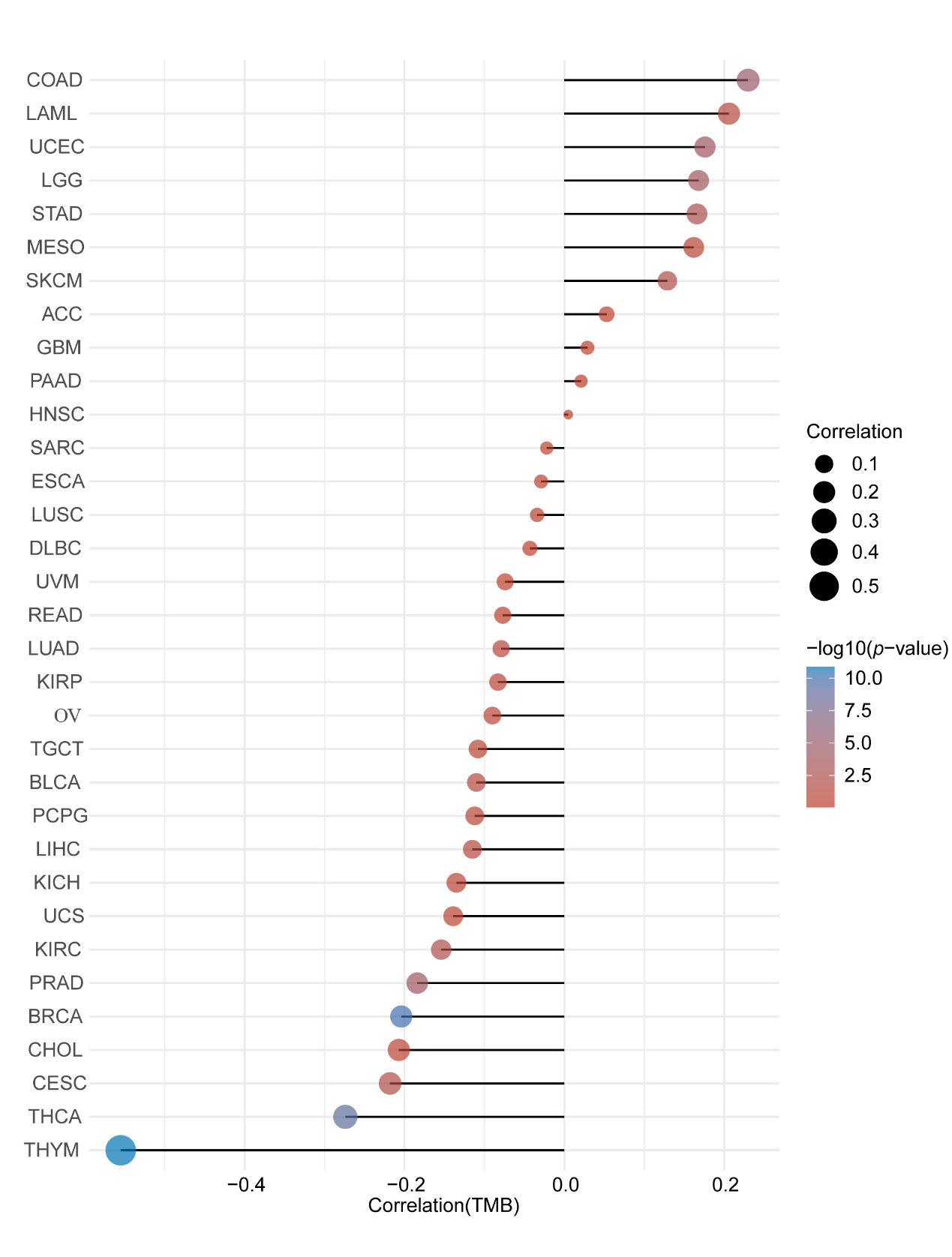

**Fig. S7**: Spearman correlation analysis of TMB and *MCPH1* gene expression. The horizontal axis in the figure represents the correlation coefficient between genes and TMB, the ordinate is different tumors, the size of the dots in the figure represents the size of the correlation coefficient, and the different colors represent the significance of the p-value. The bluer the color, the smaller the *p*-value.

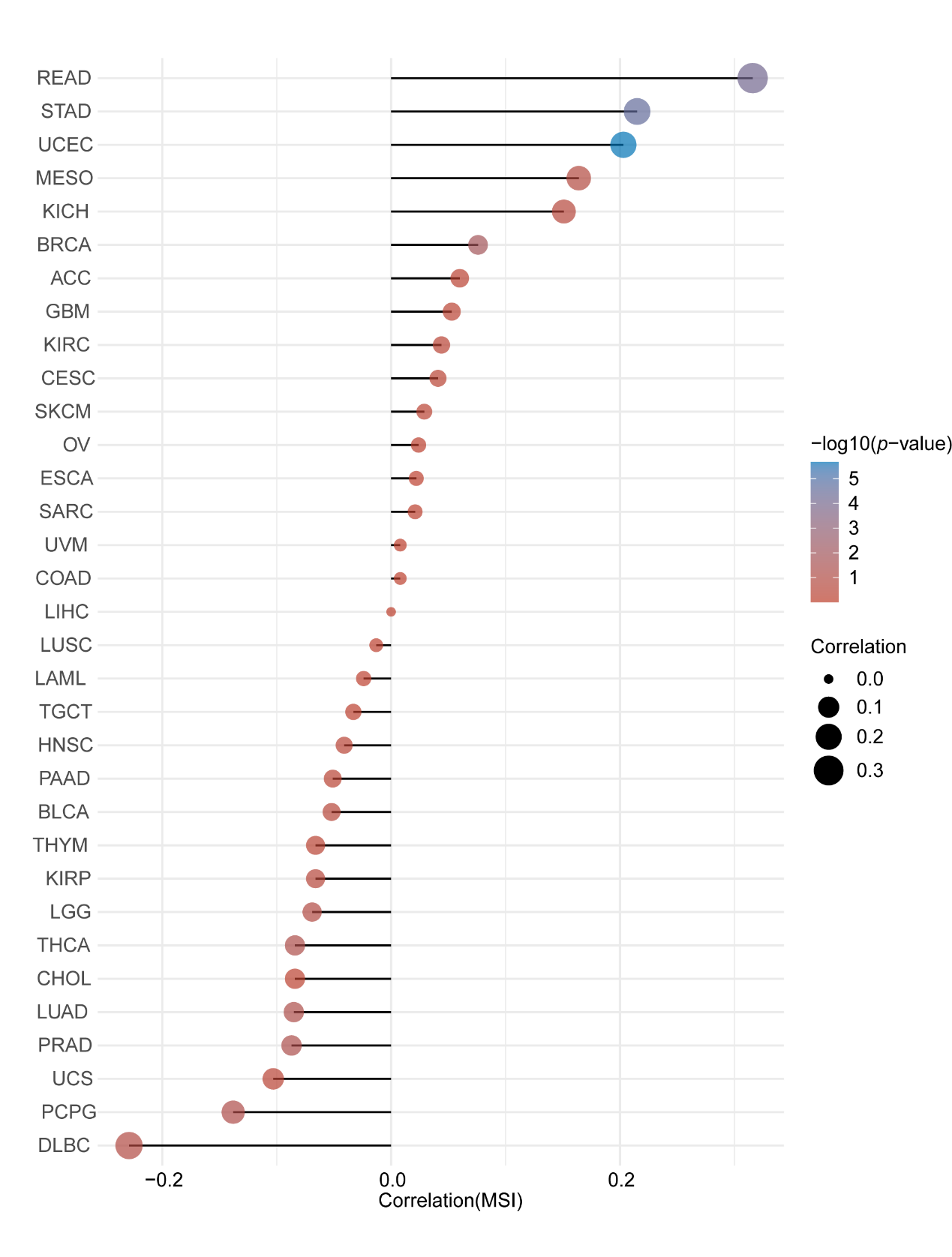

**Fig. S8**: Spearman correlation analysis of MSI and *MCPH1* gene expression. The horizontal axis in the figure represents the correlation coefficient between genes and MSI, the ordinate is different tumors, the size of the dots in the figure represents the size of the correlation coefficient, and the different colors represent the significance of the *p*-value. The bluer the color, the smaller the p-value.

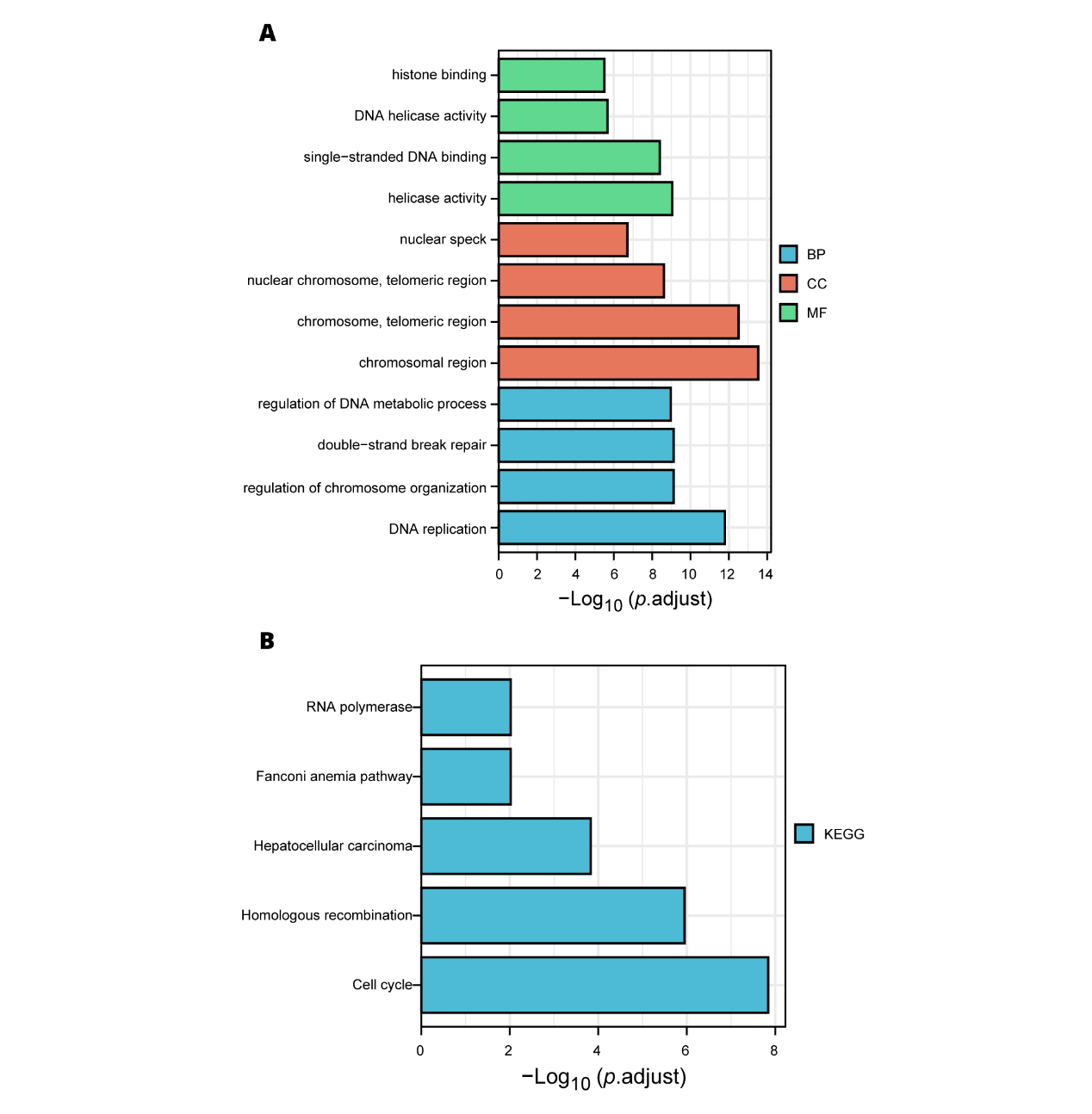

**Fig. S9**: KEGG and GO analysis of *MCPH1*. (**A**) Based on the MCPH1-binding and interacted genes, KEGG pathway analysis was performed. (**B**) The molecular function data in GO analysis.
